# Investigating the Utility of SARC-F and MSRA Questionnaires in Identifying the Risk of Sarcopenia and Falls in Juvenile Idiopathic Arthritis

**DOI:** 10.1101/2025.11.24.25338687

**Authors:** Amaani Ahmad, Sameera Oruganti, Zahra Ladan, Nicole Y Zhang, Louise Willoughby, Fatima Yousaf, Vithushi Karunakaran, Souraya Sayegh, Charalambos Hadjicharalambous, James Glanville, Madhura Castelino, Corinne Fisher, Maria Leandro, Venkat R Reddy, Debajit Sen

**Author notes:** Corresponding authors Venkat R Reddy, Debajit Sen. These authors contributed equally to this work (Amaani Ahmad, Sameera Oruganti).

## Abstract

**Introduction:** Chronic inflammation associated with ageing contributes to sarcopenia-related risk of falls and multimorbidity. The SARC-F questionnaire, based on strength, ambulation, rising from a chair, climbing stairs and falling, and MSRA (mini sarcopenia risk assessment) are validated tools to identify sarcopenia in elderly people. Here, we investigated the utility of SARC-F and MSRA in identifying the risk of sarcopenia and falls in young individuals living with juvenile idiopathic arthritis (JIA).

**Methods:** JIA patients and healthy participants were invited to complete an online survey including the SARC-F, MSRA-5, and MSRA-7 questionnaires. Additional questions explored their nutrition and exercise habits.

**Results:** We invited 101 healthy participants and 41 patients with JIA. Of the 41 JIA respondents, the median age was 23 years and 25 (61%) were female. According to the SARC-F, MSRA-5, and MSRA-7 questionnaires, 20%, 47%, and 33% were at risk of sarcopenia, respectively. Around 27% of JIA patients reported at least one fall in the past year compared to around 33% of healthy participants. However, 20% of JIA participants reported more than four falls in the past year in contrast to 4% of healthy participants. Furthermore, 44% of JIA participants reported difficulty climbing stairs and 17% reported limited ability to walk one kilometer.

**Conclusions:** Our study has demonstrated the potential utility of SARC-F and MSRA in JIA for identifying patients at risk of falls. Whether SARC-F and MSRA have the potential to identify those with sarcopenia in young people with JIA warrants further validation.

**Graphical abstract:** 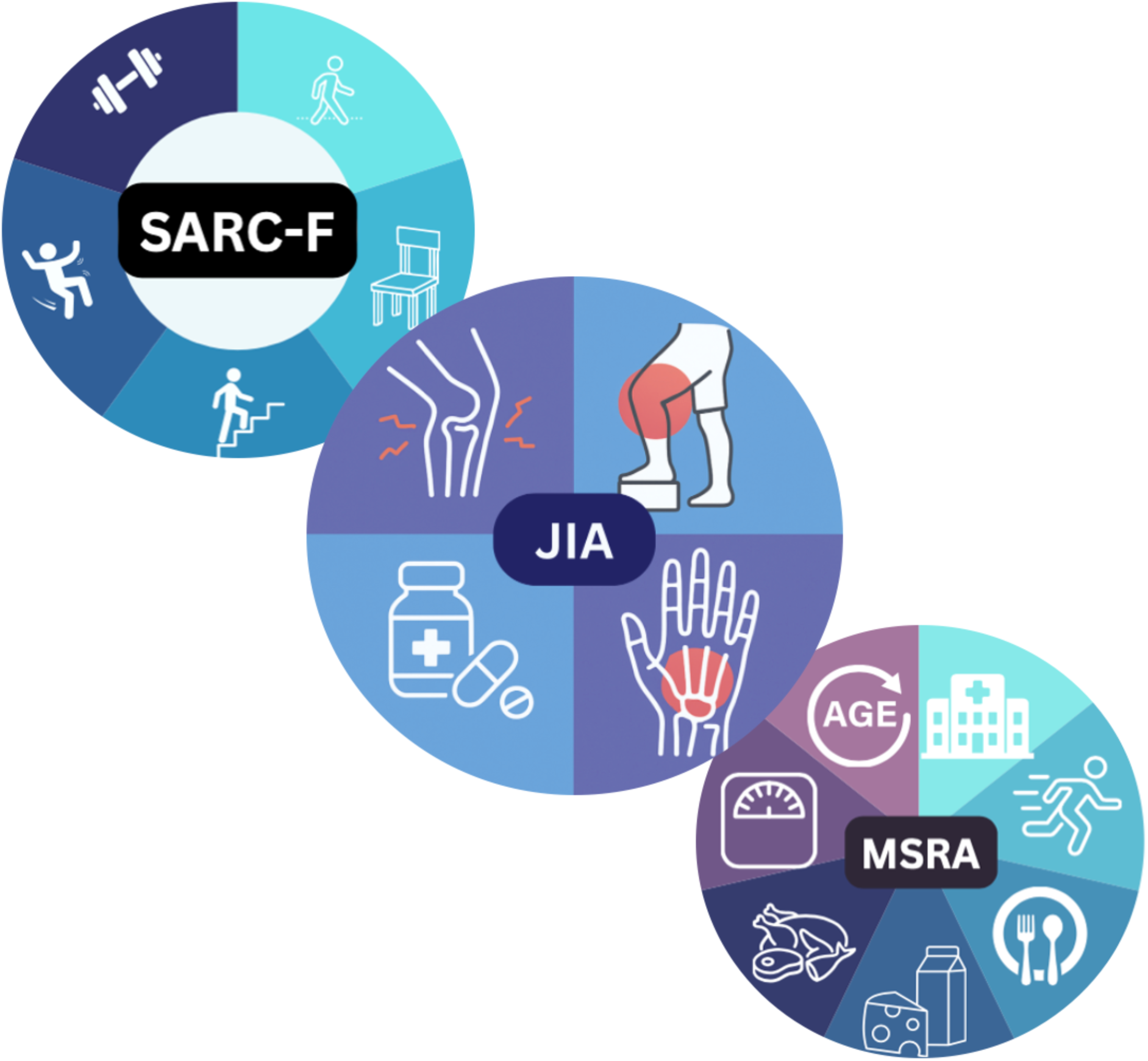

## Introduction

Chronic inflammation associated with ageing impairs muscle mass and function, known as sarcopenia (1). Sarcopenia is characterised by the gradual decline in skeletal muscle mass and function, which contributes to increased risk of falls and multimorbidity in the elderly (1, 2). Although previously considered primarily age-related, sarcopenia also occurs in chronic inflammatory disease such as rheumatoid arthritis (RA) (3, 4). Therefore, chronic inflammation is the common underlying factor driving sarcopenia in both ageing and RA.

Juvenile idiopathic arthritis (JIA) is a chronic autoimmune inflammatory disorder affecting children and adolescents (5). Similar to RA, JIA can lead to severe joint damage and significantly impact quality of life (6). Sarcopenia affects approximately 10% of individuals over the age of 65 and is associated with increased risk of falls, fractures, and overall mortality (7, 8). Limited evidence suggests that young adults with JIA have lower muscle mass than their healthy peers, related to altered bone geometry (9). Moreover, systemic glucocorticoids, a common JIA treatment, can promote muscle atrophy. Both reduced lean muscle mass and glucocorticoid exposure are independently linked to decreased bone mineral density, significantly increasing fracture risk in JIA patients (9, 10). However, we still have a limited understanding of the impact that sarcopenia has on young people with JIA who have relatively intact muscle regenerative capacity (10).

The SARC-F and Mini Sarcopenia Risk Assessment (MSRA) questionnaires were validated to identify the risk of sarcopenia and falls (11,12,14,15), as well as modifiable risk factors such as diet and exercise habits in older adults (11–14). The SARC-F questionnaire evaluates musculoskeletal (MSK) function across five key domains: muscle strength, fall frequency, walking ability, chair rise capability, and stair climbing. While SARC-F demonstrates high specificity of 99% in men and 94% in women over the age of 65, it has relatively low sensitivity (15). Nevertheless, it correlates strongly with poor muscle function, frailty, and adverse health outcomes, making it a valuable primary care screening tool (11).

The MSRA questionnaires identify at-risk individuals by assessing sarcopenia risk factors such as recurrent hospital admissions, weight loss and dietary habits. In older populations, a positive MSRA-7 score is associated with more than a fourfold increase in sarcopenia risk (12). Compared with SARC-F, MSRA questionnaires offer higher sensitivity but lower specificity, highlighting their complementary roles in screening (16, 17).

Based on these considerations, this study explores the potential utility of the SARC-F, MSRA-5, and MSRA-7 questionnaires in identifying the risk of sarcopenia and falls in individuals with JIA.

## Patients, Materials and Methods

### Participant selection

The study was conducted in accordance with NHS Research Ethics Committee approval. A cohort of healthy individuals aged over 18 years and patients with JIA satisfying the International League of Associations for Rheumatology criteria (18) and receiving care from University College London Hospital, as outlined in Supplemental Table 1, participated in the study.

### Demographic data

Demographic data including age, sex, height (cm), weight (kg) from participants were collected from electronic medical records or self-reported through online surveys.

## Sarcopenia questionnaires: SARC-F and MRSA

All participants were invited to complete a brief online questionnaire created using Microsoft Forms™. The survey comprised 29 questions, starting with the frequency of strenuous, moderate and gentle exercise. Subsequent sections covered the SARC-F questionnaire and the MSRA-5 and MSRA-7 items not available from medical records.

The SARC-F assesses five domains: strength, ability to walk unassisted across a room, ability to transfer from a chair or bed, ability to climb stairs, and fall frequency (19). In contrast, the MSRA-5 and MSRA-7 questionnaires evaluated sarcopenia risk factors such as recent hospitalisation, weight loss, and dietary habits (12). Tables 2 and 3 provide details on the SARC-F, MSRA-5, and MSRA-7 questionnaires, along with their scoring criteria.

**Table 1:**
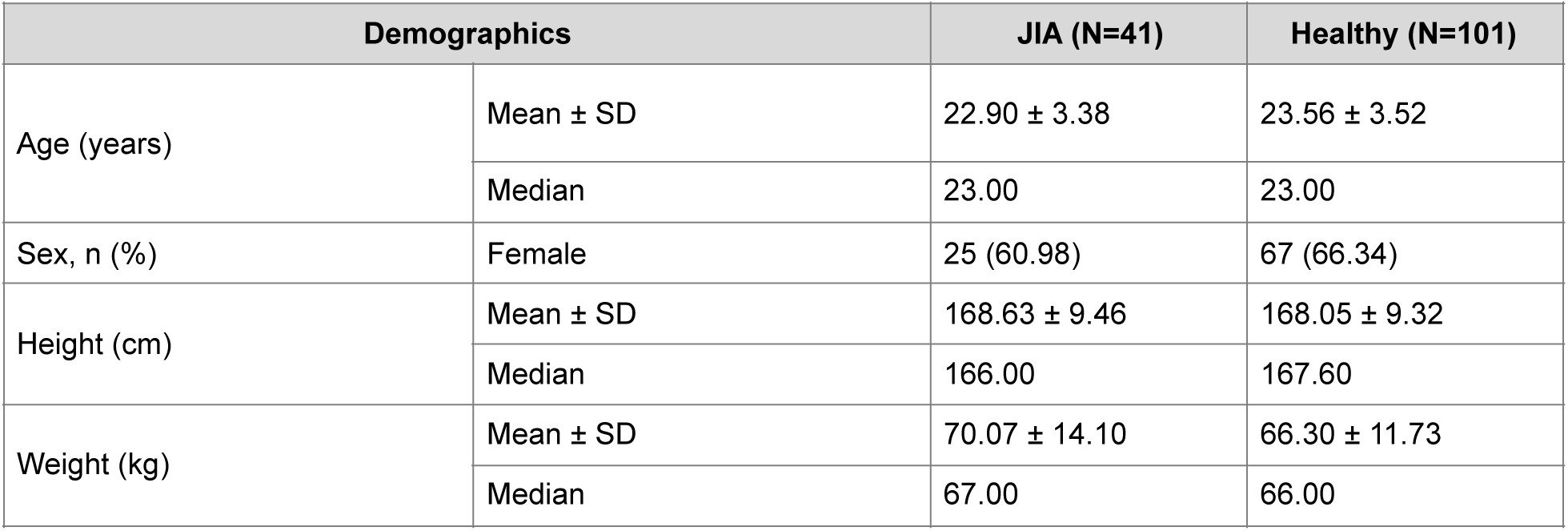
Demographic data of the JIA and Healthy cohort.

| Demographics |  | JIA (N=41) | Healthy (N=101) |
| --- | --- | --- | --- |
| Age (years) | Mean ± SD | 22.90 ± 3.38 | 23.56 ± 3.52 |
|  | Median | 23.00 | 23.00 |
| Sex, n (%) | Female | 25 (60.98) | 67 (66.34) |
| Height (cm) | Mean ± SD | 168.63 ± 9.46 | 168.05 ± 9.32 |
|  | Median | 166.00 | 167.60 |
| Weight (kg) | Mean ± SD | 70.07 ± 14.10 | 66.30 ± 11.73 |
|  | Median | 67.00 | 66.00 |

**Table 2.**
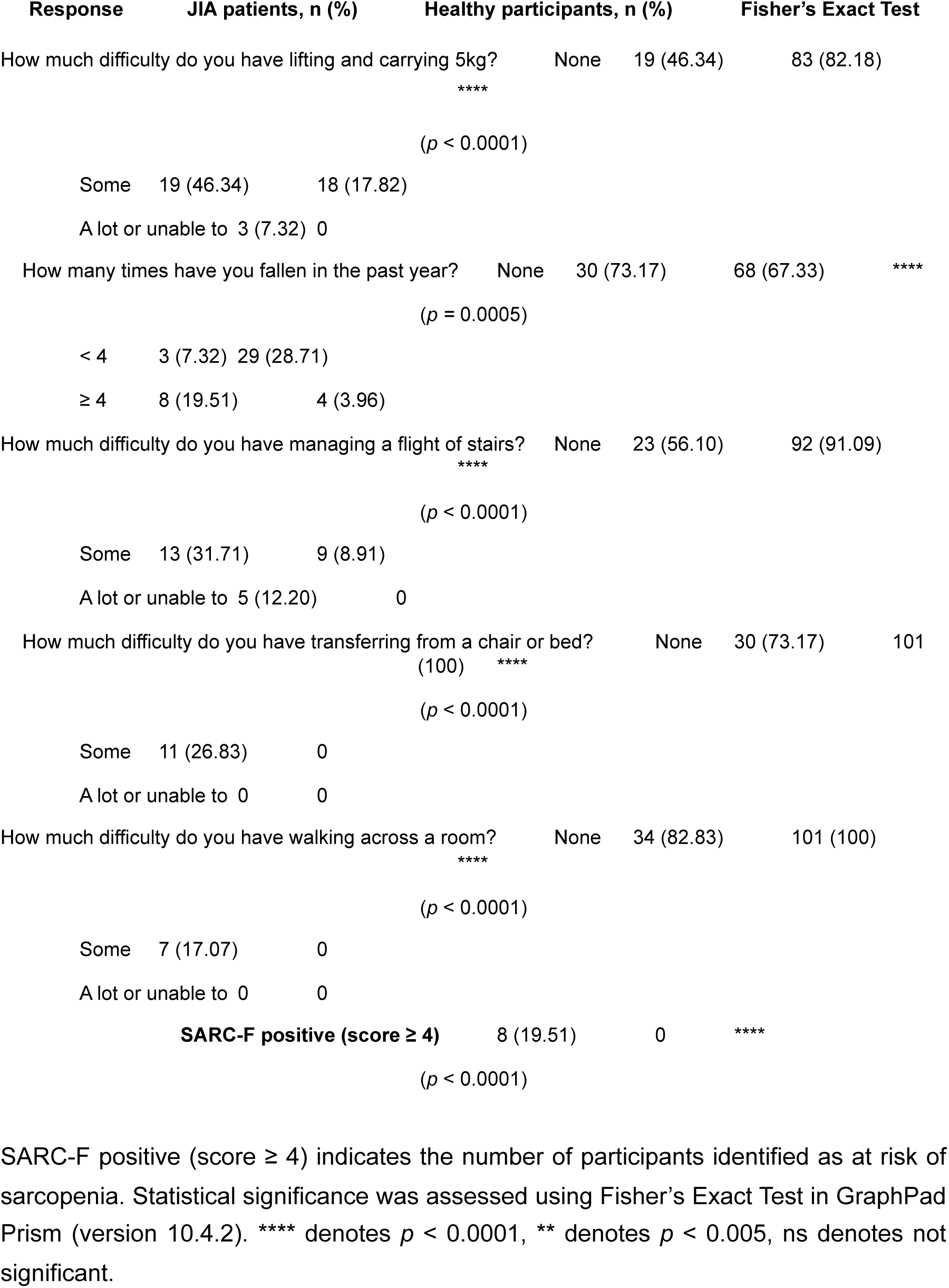
SARC-F scores and responses comparing JIA patients and healthy participants. Responses to SARC-F (5 questions) are shown for JIA patients (N = 41) and healthy participants (N = 101).

**Table 3.**
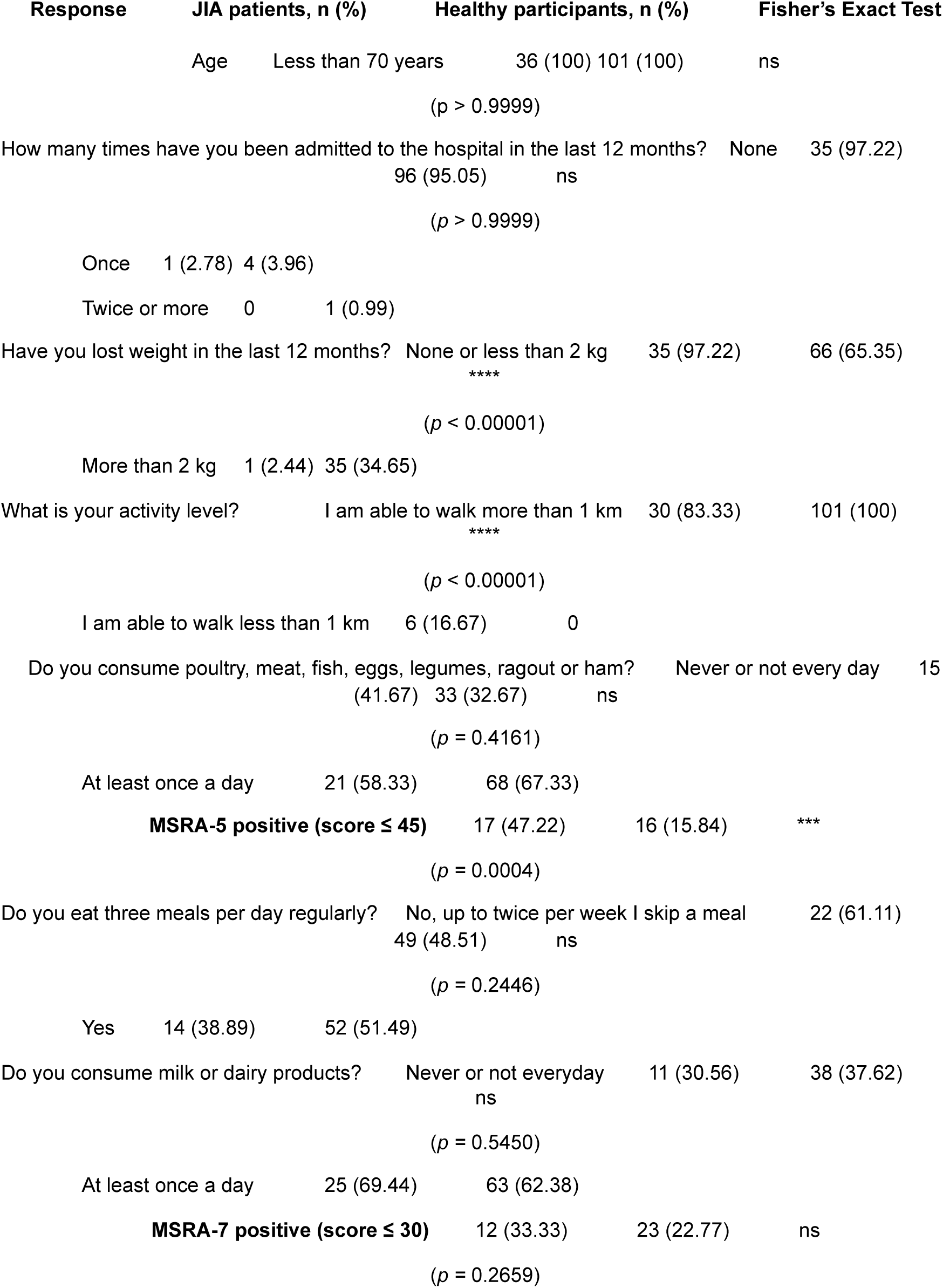
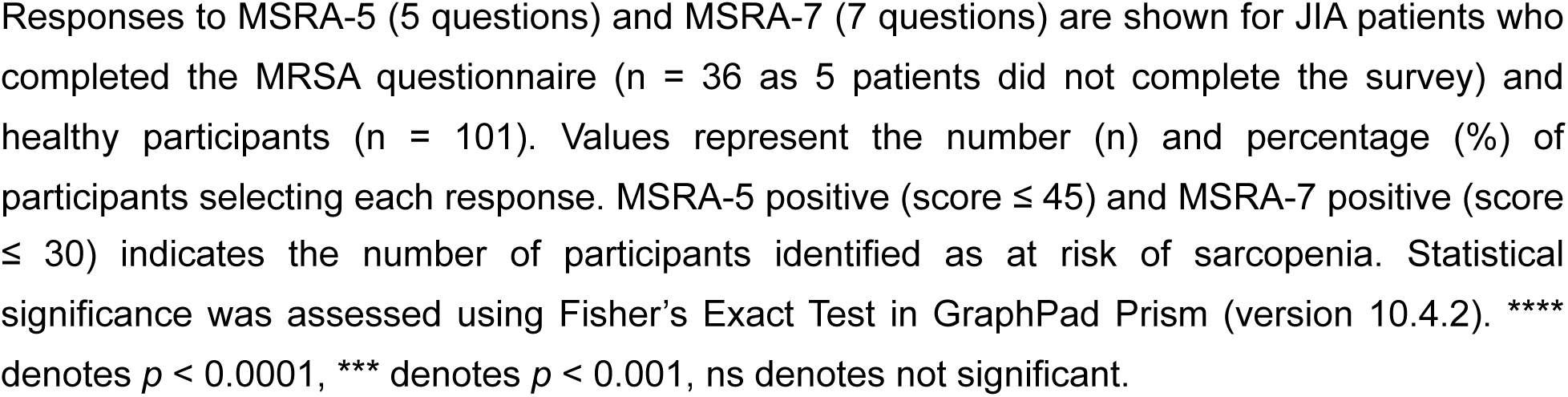
MSRA-5 and MSRA-7 scores and responses comparing JIA patients (N = 36) and healthy participants (N = 101).

| Response | JIA patients, n (%) | Healthy participants, n (%) | Fisher's Exact Test |
| --- | --- | --- | --- |
| Age | Less than 70 years | 36 (100) 101 (100) | ns |
| | | ($p > 0.9999$) | |
| How many times have you been admitted to the hospital in the last 12 months? | None | 35 (97.22) |  |
|  | 96 (95.05) | ns |  |
| | | ($p > 0.9999$) | |
| Once | 1 (2.78) | 4 (3.96) |  |
| Twice or more | 0 | 1 (0.99) |  |
| Have you lost weight in the last 12 months? | None or less than 2 kg | 35 (97.22) | 66 (65.35) |
|  | **** |  |  |
| | | ($p < 0.00001$) | |
| More than 2 kg | 1 (2.44) | 35 (34.65) |  |
| What is your activity level? | I am able to walk more than 1 km | 30 (83.33) | 101 (100) |
|  | **** |  |  |
| | | ($p < 0.00001$) | |
| I am able to walk less than 1 km | 6 (16.67) | 0 |  |
| Do you consume poultry, meat, fish, eggs, legumes, ragout or ham? | Never or not every day | 15 |  |
|  | (41.67) 33 (32.67) | ns |  |
| | | ($p = 0.4161$) | |
| At least once a day | 21 (58.33) | 68 (67.33) |  |
| **MSRA-5 positive (score $\leq 45$)** | 17 (47.22) | 16 (15.84) | *** |
| | | ($p = 0.0004$) | |
| Do you eat three meals per day regularly? | No, up to twice per week I skip a meal | 22 (61.11) |  |
|  | 49 (48.51) | ns |  |
| | | ($p = 0.2446$) | |
| Yes | 14 (38.89) | 52 (51.49) |  |
| Do you consume milk or dairy products? | Never or not everyday | 11 (30.56) | 38 (37.62) |
|  | ns |  |  |
| | | ($p = 0.5450$) | |
| At least once a day | 25 (69.44) | 63 (62.38) |  |
| **MSRA-7 positive (score $\leq 30$)** | 12 (33.33) | 23 (22.77) | ns |
| | | ($p = 0.2659$) | |

Responses to MSRA-5 (5 questions) and MSRA-7 (7 questions) are shown for JIA patients who completed the MRSA questionnaire ( $n = 36$ as 5 patients did not complete the survey) and healthy participants ( $n = 101$ ). Values represent the number ( $n$ ) and percentage (%) of participants selecting each response. MSRA-5 positive (score $\leq 45$ ) and MSRA-7 positive (score $\leq 30$ ) indicates the number of participants identified as at risk of sarcopenia. Statistical significance was assessed using Fisher's Exact Test in GraphPad Prism (version 10.4.2). \*\*\*\* denotes $p < 0.0001$ , \*\*\* denotes $p < 0.001$ , ns denotes not significant.

The final section of the questionnaire explored nutrition, including vitamin D, protein, and creatine supplementation. Participants were classified as ‘at risk’ of sarcopenia if they had a SARC-F score ≥ 4 (SARC-F positive), an MSRA-5 score ≤ 45 (MSRA-5 positive) or an MSRA-7 score ≤ 30 (MSRA-7 positive). The full questionnaire is included in the Appendix.

## Results

### Cohort demographics

A total of 142 participants completed the questionnaire, with 41 in the JIA group and 101 in the healthy cohort. Participant demographics are summarised in Table 1. The median age was 23 years in both groups. In the JIA and healthy groups, 25 (61%) and 67 (66%) of participants were female, respectively. The median weight was 67 kg for JIA participants and 66kg for the healthy cohort. Median height was 166 cm in the JIA group and 167.6 cm in the healthy cohort.

Regarding JIA subtypes, seven (17%) participants had extended oligoarthritis, polyarthritis, or enthesis-related arthritis; six (15%) participants had oligoarthritis, and two (5%) participants had systemic arthritis, psoriatic arthritis, or undifferentiated arthritis. Seven (17%) participants had a DEXA scan since their initial JIA presentation, and eight (20%) received oral or intravenous glucocorticoids in the preceding six months. These data were not collected for the healthy cohort.

#### Questionnaire responses

##### SARC-F responses

The comparative analysis of MSRA-5 and MSRA-7 responses revealed that only MSRA-5 detected a statistically significant difference in sarcopenia risk between JIA and healthy participants, while MSRA-7 did not, as shown in Table 3.

All participants were under 28 years of age, with no significant differences in age distribution between groups. Hospitalisation rates in the past year were similarly low, with 97% of JIA patients and 95% of healthy participants reporting no admissions.

Weight loss patterns differed significantly between groups. While only 2.44% of JIA patients reported losing more than 2kg in the past year, this was significantly more common among healthy participants (34.65%, p < 0.0001). Physical function assessments also revealed substantial impairment in the JIA group, with 17.07% unable to walk more than one kilometer, compared to none of the healthy participants (p < 0.0001).

Dietary patterns showed no statistically significant difference between groups. Daily consumption of protein-rich foods was reported by 58% of JIA participants, compared to 67% of healthy participants (p = 0.4161). Similarly, 69% of JIA patients and 62% of healthy participants consumed milk or dairy products at least once daily. Meal-skipping frequency was comparable, with 61% of JIA patients and 48.51% of healthy participants skipping up to two meals per week (p = 0.2446).

Figure 1 illustrates the variability in sarcopenia risk classification between JIA patients and healthy participants across screening tools. MSRA-5 identified 47% of JIA patients as at risk compared to only 16% of healthy participants (p = 0.0004). In contrast, the MSRA-7 assessment did not detect a statistically significant difference between groups, with 33% of JIA patients and 23% of healthy participants screening positive (p = 0.2659). While this could indicate similar risk profiles between groups, the lack of statistical significance may also reflect insufficient statistical power to detect a difference, given the smaller JIA sample size (n = 36).

**Figure 1:**
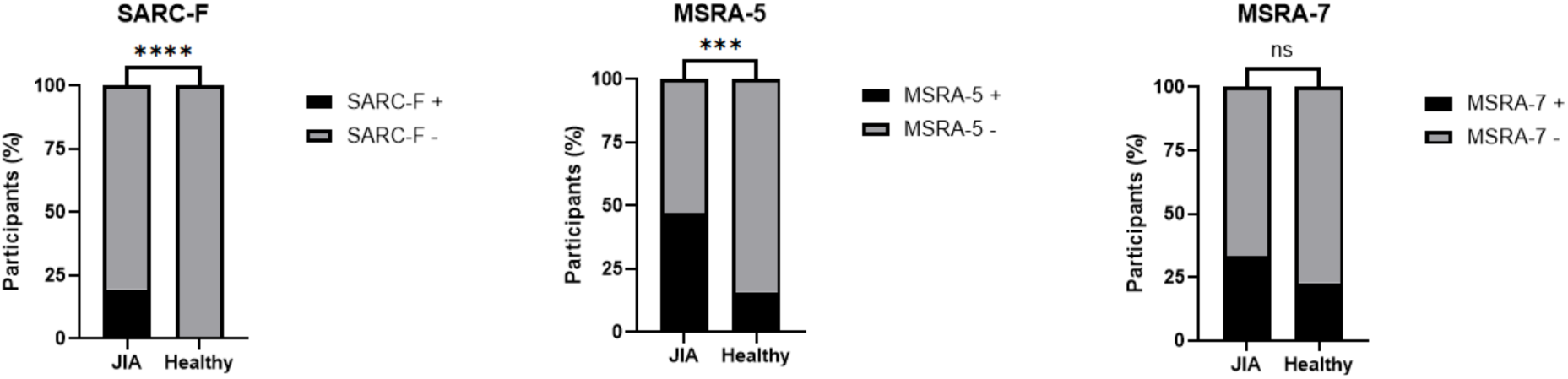
Comparison of SARC-F, MSRA-5, and MSRA-7 scores between JIA patients and healthy participants. Bar charts show the percentage of participants classified as positive (+) or negative (−) for sarcopenia based on SARC-F, MSRA-5, and MSRA-7 questionnaire scores. 19.51% of JIA patients had SARC-F+, whereas no Healthy participants had SARC-F+. The percentage of MSRA-5+ in JIA and Healthy was 47.22% and 15.84%, respectively. MSRA-7+ in JIA and Healthy was 33.33% and 22.77%, respectively. JIA patients showed significantly higher SARC-F+ (*p* < 0.0001) (N = 41) and MSRA-5+ (*p* = 0.0004) (N = 36) proportions compared to healthy participants (N = 101), with no significant difference in MSRA-7 (*p* = 0.2659). Statistical significance was assessed using Fisher’s Exact Test in GraphPad Prism (version 10.4.2). **** denotes *p* < 0.0001, *** denotes *p* < 0.001, ns denotes not significant.

These findings suggest that MSRA-5 may be more effective than MSRA-7 in discriminating between JIA and healthy populations. However, it is important to note that without gold-standard validation (such as DEXA scans, grip strength measurements, or muscle imaging), we cannot determine whether MSRA-5’s higher detection rate in JIA represents true sarcopenia risk or simply reflects disease-related functional limitations. The questionnaires were originally validated against objective sarcopenia measures in elderly populations; our study demonstrates they can detect functional differences in JIA, but their ability to accurately identify sarcopenia specifically in young adults requires validation against objective diagnostic criteria

Overall, a comparison between the three screening tools (SARC-F, MSRA-5, MSRA-7) showed that JIA patients consistently exhibited a higher risk of sarcopenia than healthy participants. However, the difference was only statistically significant in SARC-F and MSRA-5, but not in MSRA-7.

## Discussion

Our study found that the SARC-F and MSRA-5, but not MSRA-7, questionnaires can identify JIA patients at a significantly higher risk of sarcopenia and falls. Although these tools were developed to identify healthy adults / elderly populations, the findings of the study suggest their utility in young people with chronic inflammatory conditions such as JIA. While the role of chronic inflammation in sarcopenia development is well-established (4), we propose that lack of validated screening tools may contribute to inadequate recognition of sarcopenia and fall risk in younger individuals with JIA. This emphasises the importance of evaluating the utility of SARC-F and MSRA questionnaires in this context.

MSK functional impairment is well documented in ageing, and our findings also demonstrate its prevalence among young adults with JIA. In our cohort, difficulties were identified in four main domains; falls, transferring from a chair or bed, walking across a room and climbing stairs – limitations more commonly associated with sarcopenia in elderly populations. Thus, despite their young age, patients with JIA endure a significant under-recognised burden of MSK functional impairment that limits their capacity to perform daily activities.

Several factors may contribute to sarcopenia in JIA. For example, Roth *et al*.’s suggestion of reduced forearm muscle cross-sectional area in polyarticular and oligoarticular JIA patients (9), the destructive and often irreversible nature of synovial inflammation in JIA (6) and the pragmatic use of corticosteroids to control inflammation may independently increase sarcopenia risk (20). Furthermore, both JIA and sarcopenia independently elevate the risk of osteoporosis and fractures (10). Therefore, early identification of sarcopenia in this population presents a critical opportunity to mitigate these risks.

Previous validation studies assessed SARC-F accuracy against objective measures such as gait speed, grip strength and appendicular skeletal muscle mass (19). For example, in community-dwelling older adults in Hong Kong, the SARC-F demonstrated an overall accuracy ranging from 87.2% to 89.7% when compared to these physical performance metrics (15). Similarly, MSRA-5 and MSRA-7 studies reported area under the curve (AUC) values ranging from 0.681 to 0.746 for MSRA-5, and 0.713 to 0.767 for MSRA-7, indicating moderate diagnostic accuracy for MSRA-5 and low to moderate for MSRA-7 (17). However, since the mean age in these studies exceeded 75 years, the validity of these questionnaires in younger adults remains unclear (15, 17).

While our findings suggest that some individuals with JIA may have functional impairments consistent with sarcopenia risk profiles based on questionnaire results, the absence of gold-standard comparisons such as cross-sectional imaging or objective strength assessments means we cannot definitively diagnose sarcopenia. However, this study demonstrated that SARC-F and MSRA-5 successfully identify functional limitations in JIA patients that mirror those associated with sarcopenia in elderly populations. Whether these functional limitations reflect true sarcopenia (reduced muscle mass and strength) or primarily disease-specific joint pathology or a combination of both requires further investigation with objective measures.

Nonetheless, these questionnaires offer a low-cost, non-invasive and time-efficient screening method suitable for outpatient or resource-limited settings. In our study, the average completion time was 6 minutes 40 seconds for healthy participants and 7 minutes 39 seconds for those with JIA, underscoring their practicality for rapid assessment in routine care. Further validation of SARC-F and MSRA questionnaires is essential to confirm their utility in younger adults with chronic inflammatory disease.

The SARC-F and MSRA questionnaires assess sarcopenia risk through distinct yet complementary domains. SARC-F focuses on MSK function impairments such as difficulty walking, climbing stairs, or rising from a chair. In contrast, the MSRA questionnaires incorporate broader risk factors, particularly those related to nutrition and lifestyle, including protein and dairy intake, hospitalisation history, and physical activity levels. The nutritional factors in MSRA are supported by evidence from older populations, where reduced dietary protein intake correlates with accelerated muscle loss (21), and higher protein consumption (from plant and/or animal sources) is associated with slower declines in grip strength (22). Adequate calcium intake, due to its role in intracellular signalling within muscle fibres, has been inversely associated with sarcopenia risk (13). A large UK Biobank study of nearly 400,000 participants found that increased caloric, protein, and calcium intake were each associated with a reduced sarcopenia likelihood in older adults (23). However, the applicability of these findings to younger individuals with chronic inflammatory diseases such as JIA remains to be established. Younger adults possess greater physiological reserves and more efficient muscle protein synthesis (24), potentially mitigating the immediate impact of suboptimal dietary intake on muscle mass and function. The anabolic resistance seen in ageing, which requires higher protein intake to stimulate muscle synthesis, is less pronounced in younger populations (24). Consequently, while dietary inadequacies may contribute to sarcopenia risk in older adults, their direct correlation with functional impairments in younger cohorts, particularly those with JIA, warrants further investigation.

These considerations highlight the need to validate sarcopenia questionnaires in disease-specific and age-appropriate contexts, especially when interpreting components that may differ in relevance for younger populations. Our findings suggest that SARC-F and MSRA-5 may serve as clinically useful tools to identify young adults with JIA at elevated sarcopenia risk. Both questionnaires demonstrated statistically significant differences between JIA and healthy cohorts, supporting their discriminatory capacity. In contrast, the MSRA-7 did not detect significant differences, raising questions about its utility. The key distinction between MSRA-5 and MSRA-7 is the inclusion of two additional dietary-based questions in the latter: regular consumption of three meals per day and dairy intake.

The absence of significant MSRA-7 differences, despite clear MSRA-5 findings, raises questions about whether these dietary components meaningfully stratify sarcopenia risk in younger adults with inflammatory disease. In elderly populations, dietary intake is a well-established modifiable risk factor (23). However, whether the same dietary factors influence muscle health to a comparable degree in younger individuals with chronic inflammation remains unclear. Collectively, these results indicate that while SARC-F and MSRA-5 show promise, age-specific validation and potential recalibration of dietary components may be necessary to enhance utility in younger populations.

## Limitations

Although the SARC-F questionnaire is a well-established and validated tool for assessing MSK function in elderly populations, it lacks the sensitivity for detecting the functional impacts of inflammatory synovial diseases. Juvenile idiopathic arthritis (JIA) presents a wide variety of clinical manifestations, primarily affecting either the large joints of the lower limbs, as seen in juvenile oligoarthritis, or the small joints of the hands, as observed in polyarthritis. While SARC-F provides a generalised assessment of MSK function, its questions are biased towards lower-limb performance and may therefore overlook upper-limb deficits and sarcopenia.

For example, a patient with predominantly oligoarthritis affecting the knee could score in three SARC-F domains (managing stairs, transferring from a chair or bed, and falls). In contrast, a patient with polyarthritis involving only the small joints of the hands would score in just one domain (subjective strength). This discrepancy suggests that the current SARC-F may underestimate poor musculoskeletal function in the upper limbs and does not account for disease distribution. Moreover, this study did not consider disease activity.

Additionally, as domains are not described in detail in either questionnaire, there is room for misinterpretation of the question, which may lead to surprising responses. An example of this is that in the SARC-F questionnaire, 67% of healthy respondents had not fallen in the past year compared to 73% of the JIA group. Although this appears to conclude that the healthy population fell more, their definition of a fall may differ, for example, tripping over an object as opposed to falling to the ground or increased levels of physical activities such as sports. This is corroborated by the fact that even though more healthy participants fell in the past year, a greater proportion of JIA patients had more than 4 falls in the past year, 20% compared to 4%. A further example of this is that weight loss of more than 2kg was significantly more common among healthy participants (34.65%, p < 0.0001) compared to JIA patients (2.44%). As weight loss is considered to be a characteristic of sarcopenia, this is surprising. However, as the nature of weight loss was not defined, some may have interpreted it as intentional weight loss as opposed to disease-related metabolic changes and reduced activity more likely to be seen in JIA patients that preserve weight, despite muscle loss.

Since chronic inflammation is a key driver of sarcopenia, neither the SARC-F nor the MSRA questionnaires incorporate disease activity or duration of active disease. Consequently, individuals with higher disease activity and systemic inflammation, who face greater sarcopenia risk, may be underrepresented.

## Future perspectives

Although SARC-F, MSRA-5 and MSRA-7 are effective screening tools for sarcopenia, they have primarily been validated in elderly cohorts. A specialised composite questionnaire incorporating measures of systemic inflammation, disease extent and relevant SARC-F and MSRA elements should be developed to improve sarcopenia identification and prevention in JIA. Meta-analyses indicate that the HAQ questionnaire and DAS28 disease activity score correlate with sarcopenia in RA, and similar parameters should inform a composite tool for JIA (24).

## Conclusions

Sarcopenia is a disorder of skeletal muscle that affects not only older adults but also young individuals with inflammatory arthritis, such as JIA. Due to their disease, JIA patients are at an increased risk of multimorbidity, including sarcopenia. Our findings show that SARC-F effectively identified functional impairment in JIA participants, when compared with healthy controls. While both SARC-F and MSRA questionnaires are useful for assessing musculoskeletal function in JIA, further validation is required to confirm their diagnostic accuracy in younger populations. Future research should prioritise refining and validating sarcopenia screening tools for young people with JIA and other chronic inflammatory conditions.

## Data Availability

All data produced in the present study are available upon reasonable request to the authors

## Acknowledgements

VR received funding support for this work from MRC-CARP Fellowship (MR/T024968/1), Versus Arthritis and the Division of Medicine Fellowship (2024). DS and SS received funding support for this work from the MRC grant.

